# Genetic Architecture of Alcohol Use Disorder in Nepal

**DOI:** 10.64898/2025.12.13.25342207

**Authors:** Justin D. Tubbs, Yixuan He, Kristin Tsuo, Mary T. Yohannes, Younga H. Lee, Emily M. Madsen, Dirgha J. Ghimire, Sabrina Hermosilla, Tian Ge, Alicia R. Martin, Karmel W. Choi, Jordan W. Smoller, William G. Axinn

## Abstract

Alcohol use disorder (AUD) is a serious psychiatric condition associated with negative health and psychosocial consequences, comprising core symptoms of compulsive heavy drinking that impairs daily functioning. Genetic factors play a major role in AUD etiology, estimated to explain around 50% of variation in risk. Despite being a major global public health concern, the vast majority of genetic and epidemiological studies of AUD have been restricted to samples of European ancestry living in high-income countries. Using data from the Chitwan Valley Family Study, we perform the first comprehensive genetic study of AUD in a unique Nepalese sample of 10,032 individuals. We find significantly lower estimates of narrow-sense heritability (0.21, SE=0.06) compared to prior estimates from European and North American contexts. We also identified a novel genome-wide significant locus on chromosome 4 (lead SNP rs4323051, *p*=1.2e-8) in a region that determines MNS blood group status. Suggestive evidence of replication was found in a small South Asian genome-wide association study (GWAS), but not from a larger trans-ancestry GWAS. Polygenic scores for problematic alcohol use derived from an external trans-ancestry study explained 1.1% of variance in liability to AUD among males in the CVFS sample. This is the largest study of AUD in a South Asian ancestry sample and the first in a low-income country setting. Our results demonstrate the potential for careful phenotyping in ancestrally and socio-culturally diverse populations to yield novel insights into AUD etiology, while highlighting the need for larger efforts to replicate these findings and further improve cross-contextual polygenic prediction.

## Introduction

Excessive alcohol consumption can lead to numerous negative health outcomes including cancer, gastrointestinal diseases, and cognitive impairment, contributing significantly to global disability and mortality^1,2^. Moreover, harmful patterns of alcohol use also have significant social consequences including unemployment^3^ and family dysfunction^4,5^. Historically, the Diagnostic and Statistical Manual of Mental Disorders (DSM) and International Classification of Diseases have maintained independent criteria for diagnosing alcohol-related psychiatric disorders that have varied across time and place^6,7^. Broadly, alcohol use disorder (AUD) can be defined as a clinical diagnosis encompassing patterns of compulsive heavy drinking that significantly impairs daily functioning, encompassing core features across various historical nosologies.

As a complex multifactorial condition, AUD has been reliably associated with a number of risk and protective factors which vary across the lifespan. Some of the strongest psychosocial risk factors include age at first drink and externalizing personality traits for adolescent-onset AUD, childhood trauma and parental alcohol use for early middle-age-onset, and internalizing psychopathology and life events for late middle-age-onset AUD^8^. Sociocultural and economic factors such as marriage, social support, higher education, and higher socioeconomic status are associated with lower risk of AUD, especially in middle-age^8^. Although alcohol consumption behaviors and related AUD are highly culture-dependent, most studies examining risk and protective factors for AUD have been conducted in high-income countries in North America or Western Europe.

In addition to demographic and sociocultural factors, genetic risk also plays a significant role in the etiology of AUD, with additive genetic factors estimated to account for 50% of variance in liability based on twin studies from high-income North American and European settings^9^. Despite differences in prevalence, there appears to be no evidence for sex differences in the heritability of AUD^9^. Large genome-wide association study (GWAS) meta-analyses have identified over 100 genetic loci associated with AUD liability^10–12^, many of which harbor genes that encode proteins involved in alcohol metabolism, neurodevelopment, or neuronal function^13^. However, the vast majority of existing AUD GWAS rely on European ancestry samples collected in high-income countries. While recent efforts have increased representation of diverse ancestries in GWAS of AUD^10,11,14^, most samples still remain poorly reflective of global diversity across ancestry, culture, and socioeconomic status^15^.

The 12-month prevalence of AUD was estimated to be 3.7% at the most recent comprehensive global assessment in 2019, with a marginally decreasing trend since 2010^16^. However, prevalences have increased in African, Eastern Mediterranean, and South East Asian regions. Notable sex differences are apparent such that males exhibit nearly double the prevalence (4.9%) of females (2.5%) globally^16^. This difference appears even more stark in the South East Asian region, with a prevalence of 4.5% among males and 1.6% among females.

Despite its increasing prevalence and unique sex differences in these regions, AUD has been much less-studied in low- and middle-income countries (LMIC) such as Nepal, where sociocultural influences on alcohol use behaviors differ significantly from high-income countries in North America and Western Europe. The Chitwan Valley Family Study (CVFS) has been instrumental in characterizing the epidemiology of AUD in Nepal, with extensive longitudinal follow-up, careful phenotypic measurements, and low attrition^17,18^. Unique sociocultural dynamics in Nepal result in highly imbalanced rates of AUD between males and females, with women having much lower rates of ever drinking alcohol than men^19,20^. This produces a substantial difference in the lifetime prevalence of alcohol abuse disorder being 11.2% for men and 0.5% for women^19,20^. As with prior studies in high-income settings, lower levels of education and socio-economic class were associated with increased risk of lifetime AUD in Nepal^19,20^. Life events also appear to influence drinking behaviours and AUD in Nepal, with marriage exhibiting protective associations, while internal migration, exposure to armed conflict, or a transition to parenthood statistically significantly increases odds of developing AUD in males^21–23^.

Given the highly culture-dependent nature of AUD and demonstrated value of incorporating GWAS from diverse ancestries, we perform the first comprehensive genetic characterization of AUD in a unique Nepalese setting.

## Methods

### Sample

The data we use come from the CVFS, which is a whole-family longitudinal study of an ethnically diverse population in Nepal that was launched in 1995^24^. The innovative combinations of ethnography, survey data collection, and biomarker collection used in CVFS are grounded in years of prior research in Nepal and years of living among the target study population in Chitwan^24^. The sample selection took place at the neighborhood level, interviewing all households and all individuals within a selected neighborhood, and following them forward in time no matter where they moved, including throughout Nepal’s medium-intensity armed conflict^17^. The sample is fully representative of Western Chitwan and has been refreshed periodically to maintain full representation of the general population. Anyone born to these families or marrying into these families becomes part of the CVFS over the 30 years since 1995. CVFS is characterized by very high data quality, with high response rates, low longitudinal attrition, low item missing data rates, and high reliability in measures^17,18^. Portions of the World Mental Health Composite International Diagnostic Interview (WMH-CIDI 3.0) were administered to the full CVFS sample aged 15-59 in 2016-18 (with the sample representing the population of Western Chitwan in 2016)^20^. The response rate for this survey was 93%, generating 10,714 completed interviews^20^, with mental disorder measures achieving high clinical validation^25^, and 10,308 respondents providing saliva-based DNA samples. Written or verbal informed consent was obtained from all participants. All procedures were approved by the University of Michigan Institutional Review Board (HUM00104171) and by the Nepal Health Research Council.

### Phenotypic measures

#### Outcome: Alcohol Use Disorder (AUD)

Lifetime occurrence of AUD for each participant was assessed using a Nepal-specific version of the world mental health survey (WMH-CIDI) and paired life history calendar (LHC) through interviews with trained local interviewers. As detailed elsewhere, this structured diagnostic tool has undergone extensive development and validation by a multiethnic research team^26^. In particular, the LHC approach has been shown to significantly improve retrospective recall of lifetime psychiatric symptomatology ^25^. The LHC-informed WMH-CIDI used here for assessing lifetime AUD has demonstrated high clinical concordance in the Nepal population, meeting or exceeding performance of the WMH-CIDI in Western countries^25^.

#### Exposure: Childhood exposure to potentially traumatic events (PTE)

The LHC-informed WMH-CIDI included measures of exposure to PTE in childhood collected by trained interviewers. These included being badly beaten by a parent or caregiver and witnessing serious physical fights at home as a child. For this study, we combined these two questions into a single binary variable indexing childhood exposure to PTEs such that participants endorsing either question were considered exposed, and those endorsing neither were considered not exposed.

#### Covariates: Sociodemographic Variables

Sociodemographic covariates included age, genetically-inferred sex, and self-reported ethnicity. While race/ethnicity designations are complex in Nepal, the CVFS study population comprises six categories that capture key aspects of variation: Brahmin/Chhetri, Dalits, Hill Janjati, Terai Janjati, Newar, and other.

### Genomic quality control and imputation

Genotyping and quality-control (QC) have been described in a prior publication^27^. Briefly, DNA samples were genotyped at the Broad Institute using the Illumina Infinium Global Screening Array. Standardized genomic QC was performed using the GWASpy pipeline (details available here: https://github.com/atgu/GWASpy), which included filtering samples with low genotyping call rate, high inbreeding coefficient, sex mismatch, a high rate of Mendelian errors, or appearing to be duplicated samples. Phasing and imputation were performed using a new jointly called dataset of harmonized Human Genome Diversity Project (HGDP) and 1000 Genomes (1KG) Project samples^28^, providing a diverse reference panel for imputation. After genomic QC, 10,032 samples were available for analysis. Genetic principal components (PCs) were calculated in an unrelated subset of the data (N = 4,595) and remaining samples were projected onto this PC space.

### Heritability Estimation

To leverage the family-based design of CVFS, we used Genome-wide Complex Trait Analysis (GCTA)^29,30^ to estimate the narrow-sense heritability of AUD in Nepal. As outlined by Zaitlen et al.^31^, we calculated two genetic relatedness matrices using all SNPs with minor allele frequency above 0.01. A dense matrix (GRM_all_) was used to capture relatedness between all pairs of individuals, while a sparse GRM (GRM_close_) was used to model relatedness only among close family members, with elements less than 0.05 set to zero. GCTA was used to model AUD case-control status as a function of these GRMs along with fixed effects terms for age, sex, ethnicity, and the first 20 genetic PCs. An overall model, as well as two sex-specific models were fitted, providing estimates of both narrow-sense (h^2^) and SNP-based (h^2^) heritability. However, it has been noted that while GCTA produces unbiased estimates of h^2^, estimates of h^2^ from this family-based modeling approach can be highly biased. Therefore, we only report estimates of h^2^. Given that CVFS is a representative population-based sample, we used the sample prevalence of AUD to transform heritability estimates to the liability scale.

### Genome-wide Association Study

GCTA was also used to perform a mixed effect GWAS of AUD status in the full sample, as well as sex-stratified sub-samples. GRM_close_ was modeled as a random effect, with sex, age, ethnicity, and the first 20 PCs as fixed effects. Summary statistics from a prior GWAS of problematic alcohol use^10^ (a trait which includes questionnaire-based problematic drinking behavior in addition to AUD) were used to assess the replication of genome-wide significant loci.

### Polygenic Scoring

Polygenic scores (PGS) were derived using summary statistics from a large multi-ancestry GWAS of problematic alcohol use^10^. Specifically, PRS-CSx-auto^32^ was used to derive PGS weights from African, East Asian, European, and South Asian ancestry-specific summary statistics along with linkage disequilibrium statistics from respective 1000-genomes sub-populations^33^. The “meta” option in PRS-CSx was used to combine the ancestry-specific weights into a multi-ancestry inverse-variance-weighted set of PGS weights. PGS were then calculated from these weights using PLINK v2.0.0-a.6.9^34^.

### Regression Modeling

Logistic regression was used to jointly estimate the effect sizes of sociodemographic and environmental factors on risk for AUD. GCTA was used to estimate the variance explained by sociodemographic, environmental, and genetic predictors of lifetime AUD while controlling for relatedness among subjects by fitting a series of sequential linear mixed models. The baseline model included GRM_all_ and GRM_close_ as random effects and the first 20 PCs as fixed effects. Subsequent models built on this baseline model by sequentially including fixed effects for age, sex, ethnicity, PTE, PGS, and the multiplicative interaction between PTE and PGS. The variance explained (R^2^) by each predictor was calculated as the change in residual phenotypic variance after inclusion of a given fixed effect term. R^2^ estimates were subsequently transformed onto the liability scale^35^ assuming the sample prevalence as the population prevalence. In addition to a combined sex model, a male-specific model was also fitted. Female-specific models were unable to converge, likely due to the very low prevalence of AUD.

## Results

### AUD in the CVFS Sample

Among the 10,032 individuals in our sample, 6% met DSM criteria for lifetime history of AUD. Joint modeling of sociodemographic variables using logistic regression indicated that males were at significantly higher risk of AUD (OR = 24.34, 95% CI = 17.21 - 35.78, p < 2e-16), with the vast majority (95%) of cases in this sample being male. Older age at assessment was also associated with greater probability of lifetime AUD history (OR = 1.03, 95% CI = 1.02 - 1.04, p = 2.16e-15). There were also statistically significant differences in rates of lifetime AUD across ethnic groups, with Dalit (OR = 1.92, 95% CI = 1.47 - 2.49, p = 1.37e-06), Hill-Janajati (OR = 1.71, 95% CI = 1.35 - 2.16, p = 9.12e-06), and Terai-Janajati (OR = 1.92, 95% CI = 1.51 - 2.44, p = 7.73e-08) being at significantly increased risk of AUD compared to those of high-caste Brahmin/Chhetri ethnicity. Finally, self-reported exposure to a potentially traumatic event during childhood was also statistically significantly associated with greater risk of lifetime AUD (OR = 2.23, 95% CI = 1.80 - 2.76, p = 21.40e-13).

### Heritability

Using GCTA, we estimated the narrow-sense heritability of AUD in our sample to be 0.21 (SE=0.06, p = 3.8e-4), with higher heritability observed among males (0.30, SE=0.08, p = 2.2e-4). Given the small case count, female-only GCTA models were unable to converge.

### GWAS

GCTA was also used to perform GWAS in the full sample including males and females (Figure 1). GWAS analysis identified one independent locus on chromosome 4 which was significantly associated with AUD in our sample. The lead SNP (rs4323051, *p*=1.2e-8) is an intergenic SNP located within the long non-coding RNA RP11-673E1.4 and between the *GYPB* and *GYPA* genes (Figure 2). Variants in these genes have been reported in the GWAS catalog^36^ to be associated with several blood-related traits (e.g. platelet count, HbA1c levels, reticulocyte count, etc.) and also with risk for schizophrenia.

**Figure 1.**
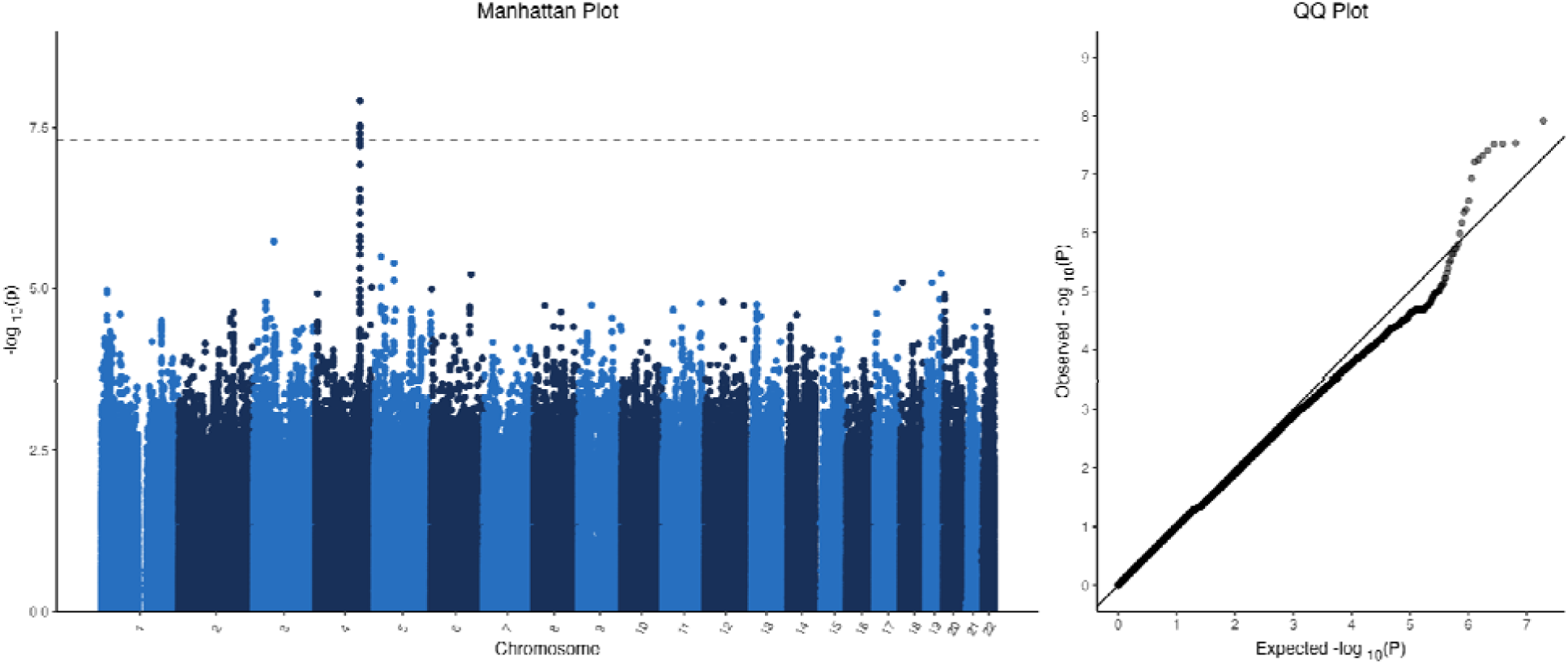
Manhattan and QQ Plots. *Note. The Manhattan plot shows single nucleotide polymorphisms (SNPs) as points, arranged by their physical position on the genome (x asis) versus the −log10(p-value) of their association with alcohol use disorder (y axis). The QQ plot compares observed −log10(p-values) on the y axis versus those expected under the null hypothesis of no association on the x axis*.

**Figure 2.**
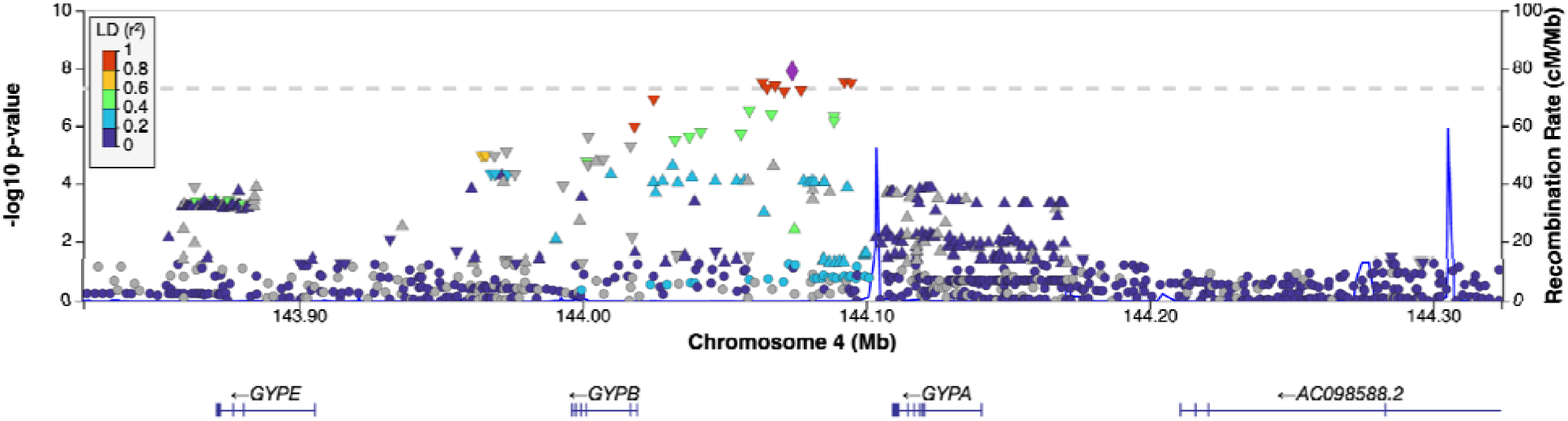
LocusZoom. *Note. This plot shows the pattern of genetic associations within the genome-wide significant locus on chromosome 4. Genomic positions are shown for build 37. Points are colored by their linkage disequilibrium (LD) with the lead variant, using LD measured in the south asian ancestry subset of the 1000 genomes reference*.

Replication of this locus was sought using summary statistics from a prior meta-analysis of problematic alcohol use in samples of South Asian ancestry from the UK Biobank and Million Veterans Program^10^ with an effective sample size of up to N=1,555. Replication results are provided in Supplementary Table 1. The lead SNP from our GWAS was only available in the UK Biobank subset of the prior GWAS, showing a consistent effect size but non-significant association p-value (P = 0.11). We also examined 22 additional non-strand-ambiguous SNPs in this locus for replication (within 500kb upstream/downstream and exhibiting within-sample correlation > 0.5 with the lead SNP). All SNPs had a consistent estimated direction of effect between the discovery and replication dataset. The lowest replication p-value was observed for SNP rs7677044 (p_discovery_=5.0e-4, p_replication_=6.1e-3). However, this locus showed no evidence of replication in a large (effective N=646,335) trans-ancestry meta analysis^10^, with only 7 out of 23 SNPs showing concordant direction of effect and the lowest replication p-value being 0.19.

### Variance Explained

The variance in risk for AUD explained by sociodemographic, environmental, and genetic variables is provided in Table 1. In the model including both males and females, sex was the strongest predictor of lifetimes AUD, explaining 25% (SE=0.7%) of the variance in liability. Age and PTE were also significant contributors, explaining 2.8% (SE=0.3%) and 2.1% (SE=0.3%) of variance in AUD liability, respectively. Ethnicity did not explain a significant proportion of variance while controlling for age, sex, and genetic PCs (which are likely to capture a high proportion of variation in self-reported ethnicity). The multi-ancestry PGS for problematic alcohol use explained a marginal, but statistically significant proportion of variance (R^2^=0.5%, SE=0.1%), while its interaction with PTE did not result in an increase in variance explained. In the male-only subsample, age (R^2^=3.9%, SE=0.4%), PTE (R^2^=3.2%, SE=0.3%), and PGS (R^2^=1.1%, SE=0.2%) explained a greater proportion of variance than in the combined sex model.

**Table 1.** Sequential Variance Explained by Sociodemographic, Environmental, and Genetic Predictors.

|  | *Male and Female Model* |  |  |  | *Male Only Model* |  |  |  |
| --- | --- | --- | --- | --- | --- | --- | --- | --- |
| *Predictor* | **R<sup>2</sup>** | **SE** | **LCI** | **UCI** | **R<sup>2</sup>** | **SE** | **LCI** | **UCI** |
| **Age** | 0.028 | 0.003 | 0.022 | 0.035 | 0.039 | 0.004 | 0.031 | 0.046 |
| **Sex** | 0.245 | 0.007 | 0.231 | 0.260 | - | - | - | - |
| **Ethnicity** | 0 | - | - | - | 0 | - | - | - |
| **PTE** | 0.021 | 0.003 | 0.016 | 0.027 | 0.032 | 0.003 | 0.026 | 0.039 |
| **PGS** | 0.005 | 0.001 | 0.003 | 0.008 | 0.011 | 0.002 | 0.007 | 0.015 |
| **PTE\*PGS** | 0 | - | - | - | 0 | - | - | - |

*Note.* We report the variance explained ( $R^2$ ) in alcohol use disorder liability by each predictor, measured as the change in residual phenotypic variance after inclusion of a given predictor. The baseline model included two genetic relatedness matrices as random effects and the first 20 PCs as fixed effects. Subsequent models built on this baseline model by sequentially including fixed effects for age, sex, ethnicity, childhood exposure to potentially traumatic events (PTE), polygenic score (PGS) for alcohol use disorder, and the multiplicative interaction between PTE and PGS. $R^2$ estimates were subsequently transformed onto the liability scale assuming the sample prevalence as the population prevalence. SE, standard error; LCI, lower 95% confidence interval; UCI upper 95% confidence interval.

## Discussion

We perform the first comprehensive characterization of genetic risk factors for AUD in Nepal, and the largest AUD GWAS to date in a South Asian ancestry sample. We estimated the narrow-sense heritability of AUD to be notably lower (0.21, SE = 0.06) than prior estimates (0.49, SE = 0.03) from studies performed in North American and European settings^9^. Our GWAS identified a novel locus on chromosome 4 with some evidence for replication in an independent sample. Finally, we found that a multi-ancestry PGS for AUD was significantly associated with risk for AUD in Nepal.

Compared to prior genetic studies of AUD, the CVFS sample is highly unique in terms of ancestry, ethnicity, and cultural context. Even within Nepal, the Chitwan Valley is notable for its ethnic diversity and dynamic changes in economics, public infrastructure, family structure, and armed conflict over the past 30 years. To our knowledge, all prior estimates of AUD heritability have come from high-income countries in Western Europe or North America. Given that alcohol consumption practices and related disorders are highly dependent on cultural context, availability, and types of alcohol, heritability estimates from a wider range of settings is vital to elucidate potential differences in etiology and develop context-appropriate intervention or prevention strategies. We find that the relative influence of genetic factors on AUD is substantially lower in Nepal compared to prior estimates^9^. Thus, social norms and practices surrounding alcohol use in Nepal appear to play a more important role in risk for AUD compared to genetic contributors. High environmental variance in the Chitwan Valley may be particularly relevant in dampening the relative contribution of genetic contributors to AUD in this unique setting.

As with heritability estimates, prior GWAS of AUD have heavily relied on samples of European ancestry from high-income countries. We report the largest GWAS to date of AUD in subjects of South Asian ancestry and the first in a LMIC setting. We identify a novel association at a locus on chromosome 4 that includes *GYPA, GYPB,* and *GYPE* genes, which encode glycophorin antigens used to define the MNS blood group system^37^. This is thought to be a complex region that has previously undergone positive selection related to malaria resistance^38^. Several early genetic linkage studies examined this locus, but findings are mixed and the resolution of these studies is coarse^39–42^. One recent epigenomic study reported a significant replicable hypomethylation of *GYPA* in brain and buccal cells of AUD cases compared to controls^43^. Our attempt at replication of the locus identified in our study yielded some supportive evidence from a small South Asian GWAS, but no evidence was found in the much larger multi-ancestry GWAS meta-analysis^10^. Further studies in South Asian ancestry samples will be necessary to provide more convincing evidence as to whether this locus may harbor an ancestry-specific association or if it is a false-positive finding. It is notable that our analysis did not identify statistically significant associations in aldehyde or alcohol dehydrogenase genes, which have strong evidence for harboring variants with protective effects on AUD, including from multi-ancestry samples^10,11,14^. Prior candidate gene studies have noted that canonical protective alleles at these loci appear at a very low frequency or are not observed in South Asian (Indian) samples^44–47^.

Multi-ancestry PGS for problematic alcohol use explained a statistically significant proportion of variance in liability to AUD in this Nepalese sample, especially among males (1.1%). In comparison, a similar PGS predicted 3.3% of variance in survey responses about problematic alcohol use in European ancestry samples^10^. Other studies have reported relatively lower variance explained by AUD PGS across various alcohol-related outcomes, ranging from 0.03% to 1.7% in European and African ancestry samples^11,48–51^.

Despite being the largest genetic study to date of AUD in the unique, diverse Nepalese population, our sample size is still modest compared to studies in other ancestries. Thus, the genome-wide significant locus reported here requires further validation in larger samples of South Asian ancestry. Nevertheless, the potential for identifying ancestry- or context-specific associations highlights the importance of increasing sample sizes of genetic studies in more socioculturally and ancestrally diverse populations. These efforts are likely to produce novel biological insights and reduce the cross-ancestry PGS prediction gap, contributing to equitable improvements in prevention, diagnosis, and treatment of AUD.

## Data Availability

We intend to make GWAS summary statistics available upon publication.

## Acknowledgements

This project was supported by the National Institute of Mental Health (JWS, WA, grant number R01MH110872), (KWC, grant number K08MH127413), (JDT grant number K01MH141330); a NARSAD Brain and Behavior Foundation Young Investigator Award (KWC, no grant number); the National Human Genome Research Institute (JDT, grant number T32HG010464); and a Eunice Kennedy Shriver National Institute of Child Health and Human Development Center Grant (DG, WA, P2CHD041028) to the Population Studies Center at the University of Michigan.

